# Rett syndrome severity estimation with the BioStamp nPoint using interactions between heart rate variability and body movement

**DOI:** 10.1101/2022.03.21.22272708

**Authors:** Pradyumna Byappanahalli Suresha, Heather O’Leary, Daniel C. Tarquinio, Jana Von Hehn, Gari D. Clifford

## Abstract

Rett syndrome, a rare genetic neurodevelopmental disorder in humans, does not have an effective cure. However, multiple therapies and medications exist to treat symptoms and improve patients’ quality of life. As research continues to discover and evaluate new medications for Rett syndrome patients, there remains a lack of objective physiological and motor activity-based (physio-motor) biomarkers that enable the measurement of the effect of these medications on the change in patients’ Rett syndrome severity. In our work, using a commercially available wearable chest patch, we recorded simultaneous electrocardiogram and three-axis acceleration from 20 patients suffering from Rett syndrome along with the corresponding Clinical Global Impression - Severity score, which measures the overall disease severity on a 7-point Likert scale. We derived physio-motor features from these recordings that captured heart rate variability, activity metrics, and the interactions between heart rate and activity. Further, we developed machine learning (ML) models to classify high-severity Rett patients from low-severity Rett patients using the derived physio-motor features. For the best-trained model, we obtained a pooled area under the receiver operating curve equal to 0.92 via a leave-one-out-patient cross-validation approach. Finally, we computed the feature popularity scores for all the trained ML models and identified physio-motor biomarkers for Rett syndrome.

## Introduction

Rett syndrome is a rare genetic neurodevelopmental disorder that occurs primarily in girls [1]. Though Rett syndrome is a clinical diagnosis, more than 95% of cases are caused by mutations in the gene encoding the methyl-CpG binding protein 2 (MECP2), a transcriptional regulator involved in chromatin remodeling and the modulation of RNA splicing [2]. Rett syndrome affects 1 in 10000 females and is characterized by a period of apparently normal postnatal development followed by developmental delay and loss of acquired skills resulting in psychomotor regression, development of stereotypical hand movements, and dysautonomia [3]. It leads to the deterioration of the autonomous nervous system, impacting breathing regularity, heart rate (HR), gut motility, and impairs motor planning and locomotion, resulting in significantly impaired mobility, no purposeful hand use, and largely absent verbal communication. There is no permanent cure for Rett syndrome in humans, and symptom management remains the standard of care [4]. When new drugs are discovered to alleviate specific Rett symptoms, clinical trials are conducted to learn about their efficacy, safety, and side effects. An essential step in measuring the efficacy of a drug or treatment method is to assess the associated benefits and risks through clinical trials. However, objective measures of symptom severity are not yet available for Rett syndrome or neurological conditions generally, and efforts to develop objective measures of autonomic symptoms could significantly enhance the ability to understand therapeutic efficacy. The key benefit we would like to see in Rett patients is improving their autonomous nervous system’s function and locomotion. Amongst various indices measured in Rett clinical trials, the Clinical Global Impression - Severity (CGI-S) is used to measure overall disease severity in Rett subjects [5, 6]. The CGI-S is a 7 − point Likert rating scale that reflects experts’ clinical judgment of the patient based on the clinician’s total experience with the Rett syndrome population. The CGI-S ranges from 1 to 7 and each score corresponds to the following patient states: (1) normal, not at all ill, (2) borderline ill, (3) mildly ill, (4) moderately ill, (5) markedly ill, (6) severely ill, (7) amongst the most extremely ill. It has been widely used as an outcome measure in Rett syndrome and other neurodevelopmental disorders such as Autism and Fragile X Syndrome [5]. In our work, we measured CGI-S in all our patients during all clinic visits included in our experiments to assess their global clinical state.

Dysautonomia or autonomic dysfunction is the abnormal function of the autonomous nervous system. It adversely affects involuntary body functions, including blood-pumping by the heart, maintaining proper blood pressure and respiration. Unfortunately, dysautonomia is a cardinal feature in Rett syndrome [7–12]. A principled approach to characterize dysautonomia is to utilize the electrocardiogram (ECG) [8, 13–15]. Researchers have used the ECG to study the variations in breathing and HR in Rett girls [16, 17]. In our work, we captured the ECG signal from girls suffering from Rett syndrome using the BioStamp^®^ nPoint wearable biosensors to capture the severity of autonomic dysfunction and, in turn, develop Rett severity classification models using ECG. Specifically, we used the heart rate variability (HRV) metrics for this purpose. The HRV is a physiological phenomenon of the variation of the time interval between the heartbeats. Typically, it is measured using a set of metrics known as the HRV metrics. We describe the HRV metrics utilized in this work in detail in the section *Materials and methods*. Dystonia is a movement disorder where a subset of muscles contract uncontrollably. The contractions cause the affected body parts to twist involuntarily, resulting in repetitive movements. In Rett syndrome, dystonia, psychomotor regression, and stereotypical hand movements are fundamental concerns and cause significant stress on patient and caregiver quality of life. We refer the readers to the following works [18–21] for a detailed explanation of dystonia in Rett syndrome. The BioStamp^®^ nPoint wearable biosensors measured the body movements of Rett patients by recording the three-axis acceleration signal via an accelerometer [22], an electromechanical sensor that senses static and dynamic forces of acceleration. To further characterize body movements, we derived rest activity metrics [23] and the cosinor rhytmometry features from the captured three-axis acceleration signals. Together, we call them actigraphy metrics. We describe these metrics in the section *Materials and methods*. Interactions between the HR and body movement were quantified using multiscale transfer entropy (MSTE) [24, 25] and multiscale network representation (MSNR) [26]. The MSTE metrics measured the information flow between two simultaneously sampled time series at multiple time scales. In MSNR, we constructed network representations of simultaneously sampled 3-dimensional time series at multiple time scales and derived network characteristics at each time scale. These network representations revealed more nuanced characteristics of the time series being analyzed.

The goal of our study was two-fold. First, we wanted to develop machine learning (ML) classification models to classify patients with low-severity Rett syndrome (CGI-S ≤ 4) from patients with high-severity Rett syndrome (CGI-S *>* 4) based on the objective measures attained from a wearable biosensor. Second, through the classification experiment, we wanted the trained models to provide us with important features (physio-motor biomarkers) that could help us distinguish the two groups. Hence, we developed Rett syndrome severity classifier models based on raw data recordings using metrics derived from the following feature sets: (1) HRV metrics, (2) Actigraphy metrics, (3) MSTE-features, and (4) MSNR-features. We used the least absolute shrinkage and selection operator (LASSO) for model training and developed logistic regression models with the L1-penalty. We developed separate models for each of the four feature sets listed above and for all possible two, three, and four combinations of these feature sets. Thus, we developed 15 binary-classification models for Rett syndrome severity classification. Finally, we listed the models’ features that were important for Rett syndrome severity classification. We illustrate the complete pipeline of our work in Fig 1.

**Fig 1.**
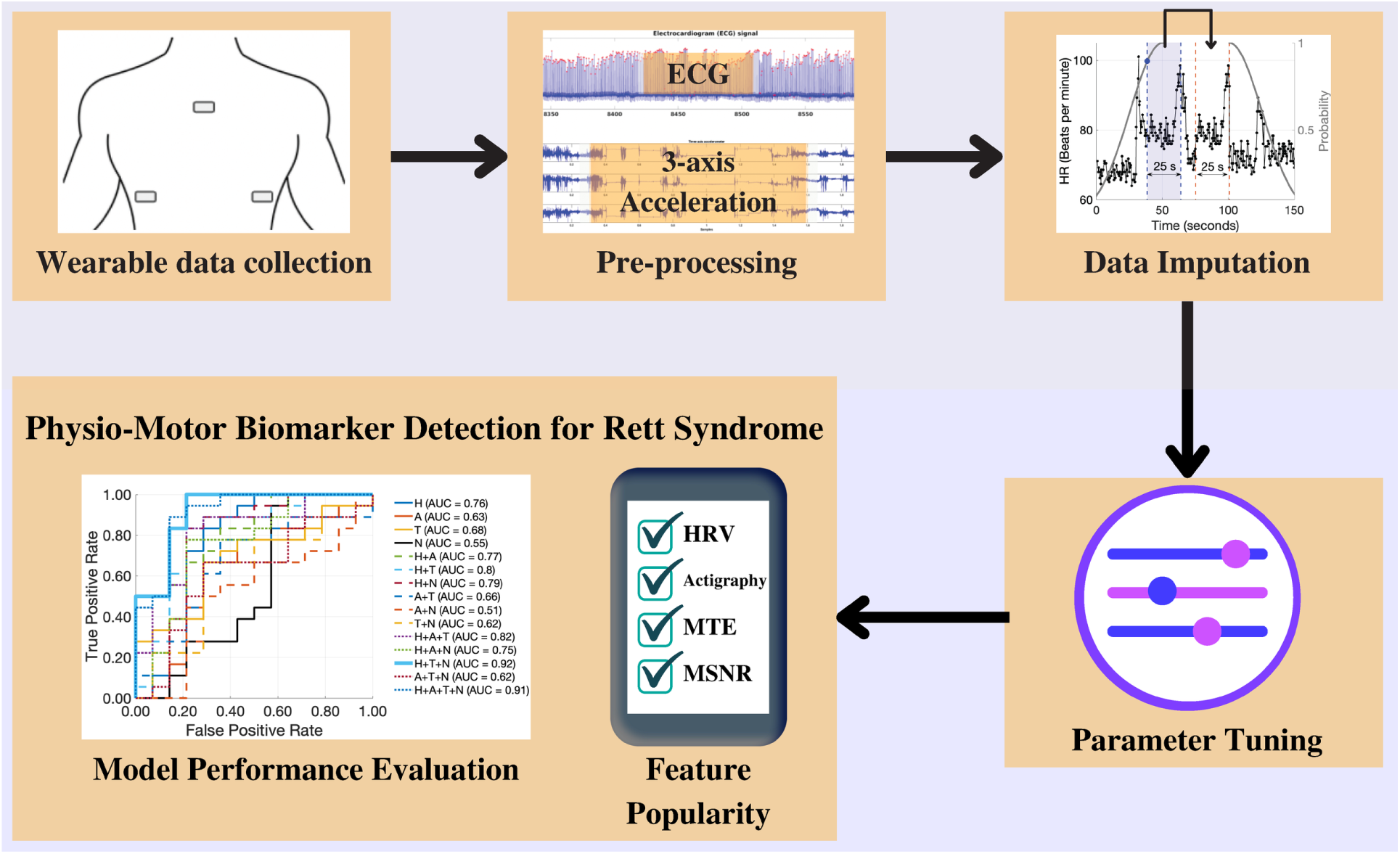
The project pipeline. We utilized the BioStamp^®^ nPoint biosensor wearables to collect simultaneous ECG and three-axis acceleration data from multiple locations on the chest. We performed data pre-processing and implemented multiple data imputation techniques to improve data quality. We trained L1-regularized logistic regression classifier models and tuned the model weights using the imputed data. Finally, we visualized model performance and computed feature popularity scores.

## Materials and methods

### Data collection

The dataset for this work was sourced from two Institutional Review Board (IRB)-approved studies: (1) The Triheptanoin-clinical trial [27] (2) The Outcome measures and biomarkers development study [28]. The data were collected between January 2016 and December 2018 - a three-year period. We used the body-worn patch BioStamp^®^ (MC10 Inc., Cambridge, MA, USA) [29] to record ECG and three-axis acceleration from all the participants. While some ECG records were captured at a sampling rate of 125Hz, others were captured at a sampling rate of 250Hz.

Concurrently, the three-axis acceleration records were captured at the sampling rates of 31.25Hz and 62.5Hz, respectively. These differences did not meaningfully influence the HRV and activity metrics we extracted [30]. We captured the ECG signal and the three-axis acceleration from the following four locations on the body: (1) Medial chest, (2) Left Hypochondrium, (3) Right Hypochondrium, and (4) Left Pectoralis. Per the protocols, all four locations were not used for all the participants, and only a subset of these locations was used for each participant. In conjunction to the signal data obtained from the biosensors, caretaker and physician surveys were conducted to obtain symptom severity for all 20 patients enrolled in the study. Specifically, the CGI-S scores were obtained through physician surveys to assign a binary label (low-severity vs. high-severity) for each patient-visit. A patient-visit was assigned to the low-severity category if the CGI-S ≤ 4 and was assigned to the high-severity category if the CGI-S *>* 4. For each patient-visit we needed two consecutive days of signal data for the feature extraction. By applying this filter, we obtained a total of 32 patient-visits with two consecutive days of signal data and the associated CGI-S label. Among the 32 patient-visits, we had 18 high-severity visits corresponding to 10 unique patients and 14 low-severity visits corresponding to 11 unique patients. One patient had both low-severity and high-severity visits. We considered each patient-visit a data point and thus had 32 data points with an associated binary label for model development and analysis.

### Study approval

This study was approved by the Emory Institutional Review Board (IRB00088492 : Outcome Measures and Biomarkers Development for Rett Syndrome and Multisite development of standardized assessments for use in clinical trials). A written informed consent was received prior to the participation from the parents of the patients.

### Missing data

Considerable amounts of missing data were present in the dataset due to the following reasons: (1) Device charging and data upload, (2) Motion artifacts, and (3) In some cases, low compliance by the caretakers. Thus, we implemented three signal imputation techniques to improve data quality and increase the amount of available data for analysis. Namely,

1. Signal quality index-based ECG data imputation.
2. Data imputation for activity counts.
3. Stochastic surrogate data imputation.

### Signal quality index-based ECG data imputation

The ECG signal recorded using the wearable patches contained sections of data corrupted by motion artifacts. To improve data quality, we sourced data from multiple sensors. As discussed earlier, we simultaneously captured ECG from up to four unique locations on the body. Thus, for a time-window t, say we had N (N ≤ 4) ECG signal snippets recorded from N locations, we chose the one signal snippet with the highest Signal Quality Index (SQI). The SQI for ECG provides the percentage of beats that match when detected by multiple annotation generators with highly differing noise responses [31]. We refer the readers to Li et. al [32] for a detailed explanation of the SQI computation algorithm. If this value was greater than 0.75, we used it in our analysis; otherwise, we discarded all ECG data for the time-window t. By switching between N signals for each time-window t to form a single 1− D ECG signal, we maximized the amount of good data available for the analysis.

### Data imputation for activity counts

When analyzing activity count signal in isolation, as per the scripts provided in the actigraphy toolbox [33], we combined the data to obtain one value per hour. When there was no data in a given hour, we imputed those samples using the Piecewise Cubic Hermite Interpolating Polynomial (PCHIP) interpolation.

### Stochastic surrogate data imputation

The computation of transfer entropy and multiscale network representation required us to impute the missing data in the 48-hour HRV features and the activity count signals. For this, we developed the surrogate data imputation method, a stochastic technique developed to impute missing data in a timeseries using data in the vicinity of the missing sections (or *gaps*). The data imputation algorithm works as follows. Given a time series (𝒮) and its timestamps (*t*), we find all the *N gaps g*[*i*] ∀*i* ∈ {1, 2, 3, …, *N* } in 𝒮 which are greater than a fixed threshold *th*_*g*_. The gaps are then sorted in increasing order. We impute the gaps with surrogate signal snippets in increasing order of the gap length as follows. We denote the gap length in seconds as *g*_*l*_, while *t*_*b*_ and *t*_*e*_ denote the time stamp where the gap begins and ends, respectively. Next, for each gap *g*[*i*] we draw a sample *x*_*r*_ from the normal distribution with mean 0 and variance equal to 1, i.e., *x*_*r*_ ∼ 𝒩 (0, 1). The normal distribution from which we picked a sample is then mapped to the timeseries 𝒮 in the following way. We map the left half of the distribution (0.5 × folded normal distribution) to 𝒮 where *t < t*_*b*_ − *g*_*l*_ and the right half of the distribution (0.5× folded normal distribution) to 𝒮 where *t > t*_*e*_. We illustrate this mapping of the Gaussian distribution onto the timeseries in Fig 2. Accordingly, we copy the signal snippet of length *g*_*l*_ starting from the point in time that corresponds to *x*_*r*_ on the timestamp signal *t*. We insert this copied signal in the gap *g*[*i*] and add a noise signal which is 5% of the sample sampled from a Gaussian distribution with mean (*μ*_*S*_) and variance 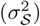 equal to the mean and variance of the signal 𝒮. Further, we update both the timeseries 𝒮 and the corresponding timestamps t. This procedure is repeated iteratively until all gaps {*g*[*i*]} in 𝒮 greater than *th*_*g*_ are imputed.

**Fig 2.**
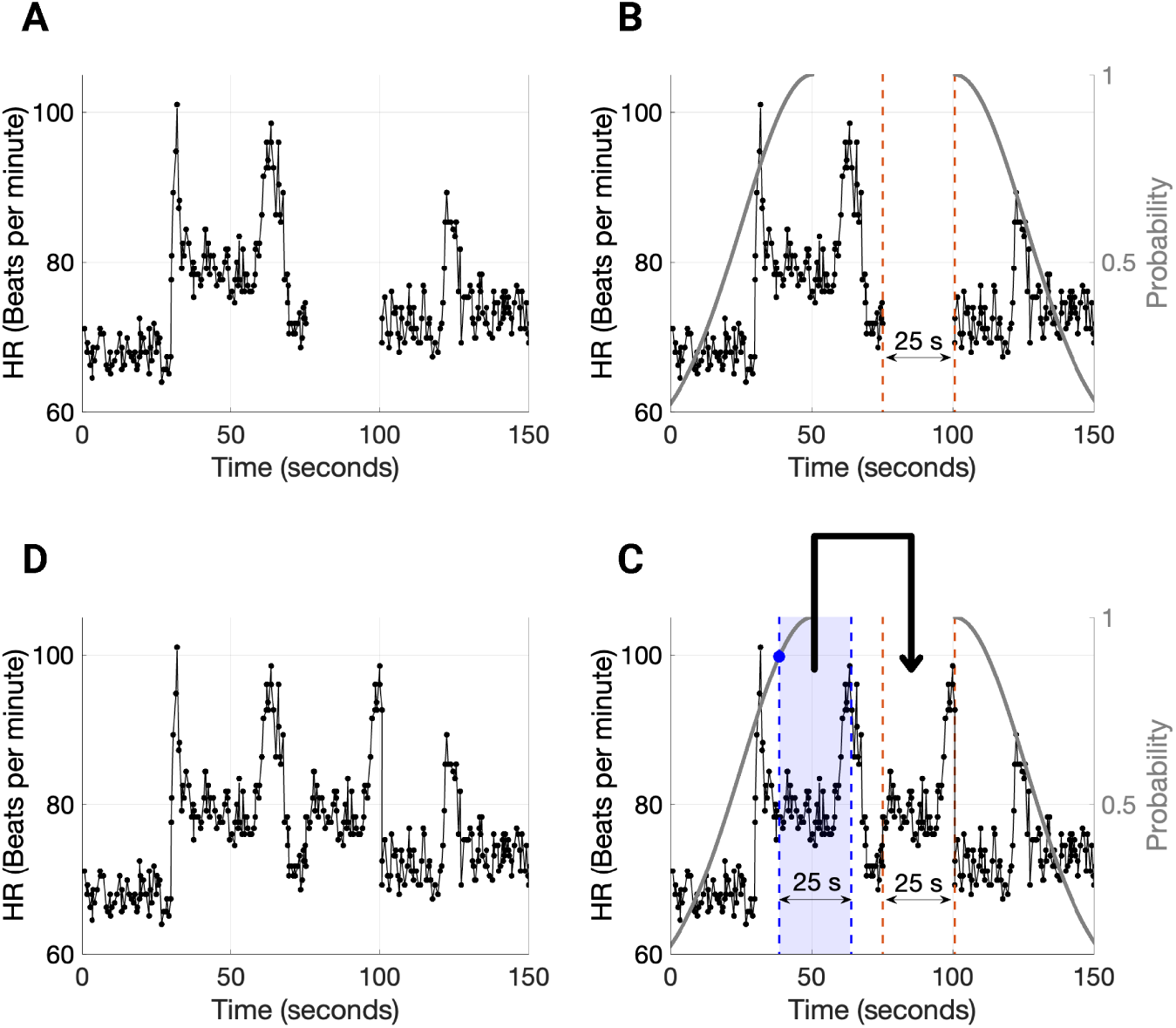
Stochastic surrogate data imputation technique fills gaps in heart rate (HR) and activity count signals by choosing a contiguous HR segment in the gap’s neighborhood of the same length as the gap. (A) We illustrate an example HR signal with missing data. (B) The HR signal contains a 25-second-long gap that has been identified. Based on the boundaries and length of the gap, two folded normal distributions are constructed for stochastically choosing a 25-second-long HR segment. (C) We perform a coin toss experiment, and based on the outcome, we sample from one of the two folded normal distributions and accordingly select a contiguous 25-second-long HR segment and copy it over to the gap. Further, we add a small noise to this imputed signal (we do not show it in the figure for convenience). (D) The imputed HR signal with no gaps.

### Feature extraction

We extracted HRV metrics from the ECG signals and actigraphy features from the three-axis accelerometer signals in the dataset. The HRV metrics were extracted using the open-source PhysioNet Cardiovascular Signal Toolbox provided by Vest et al. [31]. We extracted 24 distinct HRV metrics, including time-domain measures, frequency-domain measures, entropy measures, phase rectified signal averaging (PRSA) measures, and other non-linear metrics. We used the default window length settings provided in the toolbox and thus used a 300-second-long feature extraction window with a 30–second shift. We used the SQI based ECG data imputation to maximize the amount of good ECG data. For a given patient-visit, we computed the mean and variance of each HRV metric between the times 10 PM and 10 AM. We chose this period to include ECG data during sleep and discard the noisier signal recorded during daytime and evenings. Thus, each HRV metric provided two features resulting in 48 features from 24 HRV metrics.

We extracted the actigraphy features from the z-axis of the acceleration signal from the right hypochondrium using the open-source Actigraphy Toolbox [33]. We converted the acceleration signal to activity counts using the Oakley method described by Borazio et al. [34]. First, we used Oakley’s method for converting accelerometer signals to activity count. We then extracted the following eight features using the toolbox: (1) Interday stability, (2) Intraday variability, (3) Least active 5 hours, (4) Most active 10 hours, (5) Rest activity, (6) Mesor, (7) Amplitude, and (8) Acrophase. The last three features were based on Cosinor Rhythmometry. The Actigraphy features needed two consecutive 24− hour periods (midnight to midnight) of data for feature computation. Thus, we identified the best two consecutive 24-hour periods with the least missing data for each patient-visit. If both days did not have at least 12 − hours of acceleration data per day, those patient-visits were discarded. To impute missing data, we used the PCHIP interpolation.

Finally, we computed MTE and MSNR features using 2 − day consecutive HR and activity count signals. We utilized the *Stochastic surrogate data imputation* technique to impute missing data. We processed the HR signal, deceleration capacity (DC) of the RR-interval [35] signal, and the activity count (Act) signal for computing the features. Transfer entropy depicted as *TE*_*X*→*Y*_ is a measure of directional coupling between two concurrently sampled timeseries *X* = {*x*_1_, *x*_2_, …, *x*_*N*_ } and *Y* = {*y*_1_, *y*_2_, …, *y*_*N*_ }.

Formally, *TE*_*X*→*Y*_ is a reduction in uncertainty, given by the conditional entropy of *y*_*i*_ given its past values minus the conditional entropy of *y*_*i*_ given both its past values and past values of the other variable 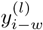:

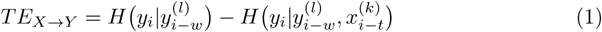

where *i* indicates a given point in time, *t* and *w* are the time lags in *X* and *Y* respectively, and *k* and *l* are the block lengths of past values in X and Y respectively. The *k* and *l* were both set to 1 to improve computational speed, and *t* and *w* were both set to 1 under the assumption that the maximum auto-transfer of information occurs from the data point in *X* immediately before the target value in *Y*, and vice versa. These choices of *k* = *l* = *t* = *w* = 1 are appropriate in biomedical experiments as the absolute values of auto-correlation functions tend to decrease monotonically as time lag increases [25]. In our experiments, we computed the TE between the following signals: (1) HR-DC, (2) HR-Act, (3) DC-Act, (4) DC-HR, (5) Act-HR, and (6) Act-DC. We computed these TE values at scales 1 to 10 using the coarse-graining algorithm [36] to obtain Multiscale Transfer Entropy (MSTE) features. The probability densities for the estimation of MTE were estimated using the Darbellay-Vajda (D-V) adaptive partitioning algorithm [25, 26, 37]. Further, we computed 3D D-V partitioning using the HR-DC-Act signals and computed multiscale network representation (MSNR) features. The network representation features included the following 11 metrics: (1) Number of nodes (total number of nodes in the network), (2) Average degree (the average value of the degree of all nodes in the network, where the degree of a node is defined as the total number of its neighboring edges), (3) Number of loops (the total number of independent loops in the network, also known as the “cyclomatic number” or the number of edges that need to be removed so that the network cannot have cycles), (4) LOOP3 (the total number of loops of size 3 in the network), (5) LOOP4 (the total number of loops of size 4 in the network), (6) Average clustering coefficient - 1 (number of LOOP3s divided by the number of connected triples in the network), (7) Average clustering coefficient - 2 (the clustering coefficient *c*(*u*) for node *u* can be defined as the ratio of the number of actual edges between the neighbors of *u* to the number of possible edges between them, and the average clustering coefficient *C*(*G*) of a network is the average of *c*(*u*) taken over all the nodes in the network), (8) Graph radius (the eccentricity of a node u is defined as *e*(*u*) = *max {d*(*u, v*) : *v∈V*}, where the distance *d*(*u, v*) is the length of the shortest path from u to v, and V is the set of all nodes; the graph radius is the minimum of eccentricities over all nodes in the network), (9) Spectral radius (the largest magnitude eigenvalue of the adjacency matrix of the network), (10) Trace (sum of the eigenvalues of the adjacency matrix, i.e.,Σ*λ*), and (11) Energy (squared sum of the eigenvalues of the adjacency matrix A. More formally, the energy of a network G is: 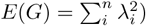. We computed the above-described MSNR features at scales 1 to 10 using the coarse-graining technique [36]. Using the surrogate data imputation technique, we obtained 100 imputations for each patient-visit. Thus, we generated 3200 datapoints. We computed the MTE and MSNR features for all the 3200 datapoints and then computed the mean and variance over 100 imputations for each patient-visit. In the end, we obtained 32 vectors of length 120 (60 mean values and 60 variance values) as the MTE features and obtained 32 vectors of length 220 (110 mean values and 110 variance values) as the MSNR features. Thus, we obtained 48 HRV features, 8 Actigraphy features, 120 MTE features, and 220 MSNR features for each of the 32 patient visits.

### Rett syndrome severity classification experiments

We developed separate models for each feature sub-group and combinations of feature sub-groups to obtain 15 classifiers corresponding to the following feature combinations: (1) HRV, (2) Actigraphy, (3) MSTE, (4) MSNR, (5) HRV + Actigraphy, (6) HRV + MSTE, (7) HRV + MSNR, (8) Actigraphy + MSTE, (9) Actigraphy + MSNR, (10) MSTE + MSNR, (11) HRV + Actigraphy + MSTE, (12) HRV + Actigraphy + MSNR, (13) HRV + MTE + MSNR, (14) Actigraphy + MTE + MSNR, and (15) HRV + Actigraphy + MTE + MSNR. We used the LASSO based logistic regression classifier to assess the performance of different feature combinations. The models were assessed via a leave-one-patient out cross-validation experiment. The hyperparameter tuning was performed using a within-training-three-fold cross-validation. We measured the classification performance using the metric - area under the receiver operating curve (AUC). Using this metric, we compared the 15 different classifiers and assessed their classification performance. For each feature combination, the entire classification experiment was repeated five times with different random seeds to account for variability in model coefficients that arose during hyperparameter tuning (the within-training-three-fold data split changed each time). The model’s outputs for the five repetitions of the experiment were combined by computing the median value for the classification probability output. The final AUC for each feature combination was determined by comparing this median output against the ground truth labels.

### Feature popularity score

Apart from measuring classification performance, we computed a novel feature popularity score for the features used in classification, which allowed us to measure feature importance and compare features. Since all classification experiments comprised 20 patients, the leave-one-patient out cross-validation approach produced 20 models. As described previously we repeated the classification experiment five times for a given feature combination, resulting in 5 × 20 = 100 classifier models per feature combination.

In these 100 models, for each feature *f*, we counted the number of models in which it had a non-zero LASSO coefficient. Then, the feature popularity score (*ρ*) for the feature *f* was computed as:

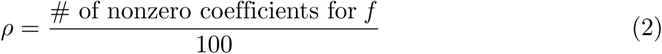

This metric measured the popularity of the feature; the greater number of times the feature was picked by the LASSO-based classifiers, the more popular the feature was.

## Results

The HRV, breathing, and physical activity are thought to influence Rett syndrome severity, but the underlying correlations are yet to be measured. Our experiments investigated the effect of HRV-metrics, actigraphy, MSTE, and MSNR features on Rett syndrome severity by assessing 15 binary classification models (Fig 3A) described in the section *Rett syndrome severity classification experiments*. For this, as described in the section *Data collection*, we utilized a cohort of 20 patients with Rett syndrome, of which 10 patients had low-severity patient-visits, nine patients had high-severity patient-visits and one individual had both low-severity and high-severity patient-visits. We illustrate this in Fig 3B.

**Fig 3.**
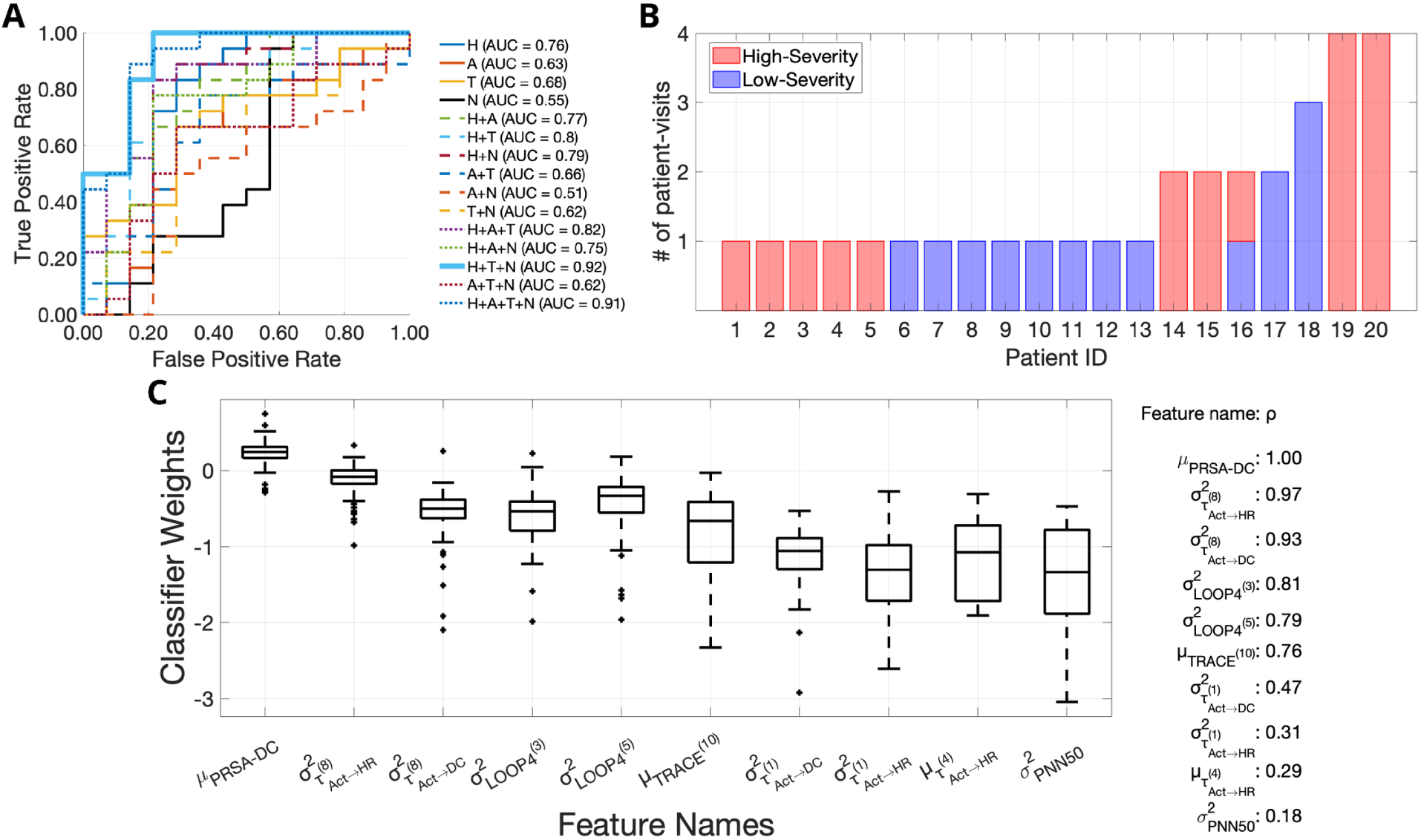
The feature combination of heart rate variability (HRV) metrics, multiscale transfer entropy, and multiscale network representation achieves the highest area under the receiver operating curve (AUC), equal to 0.92. (A) The ROCs for the 15 different Rett severity classifiers are provided. Each classifier uses a different subset-combination of the four feature sets, namely: (1) HRV metrics (H), (2) Actigraphy metrics (A), (3) Multiscale transfer entropy features (T), (4) Multiscale network representation features (N). The combination of H+T+N performed the best with a leave one-patient out cross-validation pooled-AUC equal to 0.92. The individual ROCs corresponding to the individual classifiers are shown using a combination of line styles and colors. The figure legend shows the pooled-AUC values for each feature combination. (B) A depiction of the number of patient-visits for each of the 20 patients showing the low-severity and high-severity patient-visits in different colors. (C) The top 10 most popular features used by the best classifier (H+T+N) for Rett severity classification are shown here, along with the corresponding feature coefficients. The mean deceleration capacity (*μ*_*PRSA*−*DC*_) is the most popular feature with a feature popularity score of 1 followed by 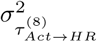 and 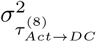 with feature popularity scores 0.97 and 0.93, respectively. In the top-10 most popular features for the H+T+N feature combination, two features were HRV-metrics (*μ*_*PRSA*−*DC*_ and 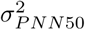), three features were MSNR-features (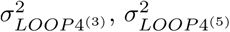 and 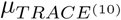) and the rest five were MSTE-features.

### Binary Classification Performance

The best binary Rett severity classifier used the feature combination of HRV-metrics, MSTE-features, and MSNR-features, and obtained a pooled-AUC equal to 0.92. When we used the four feature sub-groups separately for classification, HRV-metrics performed the best with a pooled-AUC equal to 0.76. This was followed by MSTE-features (AUC = 0.68), Actigraphy-metrics (AUC = 0.63) and MSNR-features (AUC = 0.55) respectively. When we used 2-combinations of feature sets, the feature combination of HRV and MSTE performed the best with an AUC = 0.80. This was followed by the following 2-combinations of feature sets: (1) HRV + MSNR (AUC = 0.79), (2) HRV + Actigraphy (AUC = 0.77), (3) Actigraphy + MSTE (AUC = 0.66), (4) MSTE + MSNR (AUC = 0.62), and (5) Actigraphy + MSNR (AUC = 0.51). When we used 3–combinations of feature sets, we obtained the following descending order of classification performances: (1) HRV + MSTE + MSNR (AUC = 0.92), (2) HRV + Actigraphy + MSTE (AUC = 0.82), (3) HRV + Actigraphy + MSNR (AUC = 0.75), (4) Actigraphy + MSTE + MSNR (AUC = 0.62). Finally, when all feature subsets were used together, we obtained an AUC = 0.91, only second to the combination of HRV-metrics, MSTE-features and MTNR-features (AUC = 0.92).

### Feature popularity

The mean deceleration capacity (*μ*_*PRSA*−*DC*_) was the most popular feature for Rett syndrome severity classification. Using the novel formula described in the section *Feature popularity score*, we extracted the top-10 most popular features utilized by the best classifier (HRV, MSTE, and MSNR) and the distribution of their corresponding weights (Fig 3C). The feature *μ*_*PRSA*−*DC*_ came out on top with a feature popularity score *ρ* = 1.00. It was followed by the following 9 features: (1) Variance (across all surrogate representations) of transfer entropy from the activity counts signal to the HR signal at the 8^*th*^ coarse-graining scale 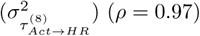, (2) Variance (across all surrogate representations) of transfer entropy from the activity counts signal to the deceleration capacity signal at the 8^*th*^ coarse-graining scale 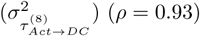, (3) Variance (across all surrogate representations) of the number of 4 − loops in the network representation of the tuple - (HR signal, activity count signal, DC signal) at the 3^*rd*^ coarse-graining scale 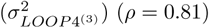, (4) Variance (across all surrogate representations) of the number of 4−loops in the network representation of the tuple - (HR signal, activity count signal, DC signal) at the 5^*th*^ coarse-graining scale 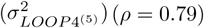, (5) Mean (across all surrogate representations) of the trace of the adjacency matrix of the network representation of the tuple - (HR signal, activity count signal, DC signal) at the 10^*th*^ coarse-graining scale 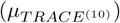 (*ρ* = 0.76), (6) Variance (across all surrogate representations) of transfer entropy from the activity counts signal to the deceleration capacity signal at the 1^*st*^ coarse-graining scale 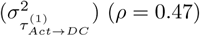, (7) Variance (across all surrogate representations) of transfer entropy from the activity counts signal to the HR signal at the 1^*st*^ coarse-graining scale 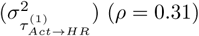, (8) Mean (across all surrogate representations) of transfer entropy from the activity counts signal to the HR signal at the 4^*th*^ coarse-graining scale 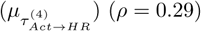, (9) Variance of the 5-minute PNN50 measure i.e., the average number of pairs of adjacent beat-to-beat intervals differing by more than 50 ms, between the times 10 PM and 10 AM 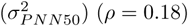. The feature weight for (*μ*_*PRSA*−*DC*_) was greater than zero for 93% of the time, suggesting an inverse relationship (due to the negative-log relationship between features and odds) between the feature values and the Rett disease severity (i.e., a higher values of *μ*_*PRSA*−*DC*_ corresponded to a higher chance of low-severity Rett syndrome). It conformed with our feature values as shown in Fig 4B. In supporting information S1 Fig we illustrate the top 5 most popular features for each of the 15 classifiers. Whenever HRV-metrics were used for classification (8 of the 15 instances), *μ*_*PRSA*−*DC*_ was the most important feature with a consistent feature popularity score of 1. When only Actigraphy-metrics were used for classification, the amplitude of the circadian rhythm measured using the cosinor rhytmometry was the most popular feature with a feature importance score of 0.94. When we used MSTE-features alone, 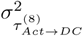 was the most popular feature with a feature popularity score of 0.94. Finally, when we used the MSNR features alone, 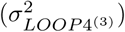 was the most popular feature with a feature popularity score of 0.90. Interestingly, the top features from the individual feature set models for HRV-metrics (*μ*_*PRSA*−*DC*_), MSTE-features 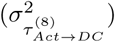, and MSNR-features 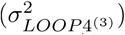 were all featured as one among the top−5 most popular features in the best model (AUC = 0.92) that used the feature combination of HRV-metrics, MSTE-features, and MSNR-features. For the classifier that used all available features (HRV + Actigraphy + MSTE + MSNR), the following metrics featured as the top-5 most popular features: (1) *μ*_*PRSA*−*DC*_ (*ρ* = 1.00), (2) 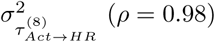, (3) 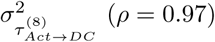, (4) Interdaily Stability (*ρ* = 0.82); (5) 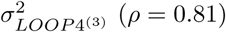

**Fig 4.**
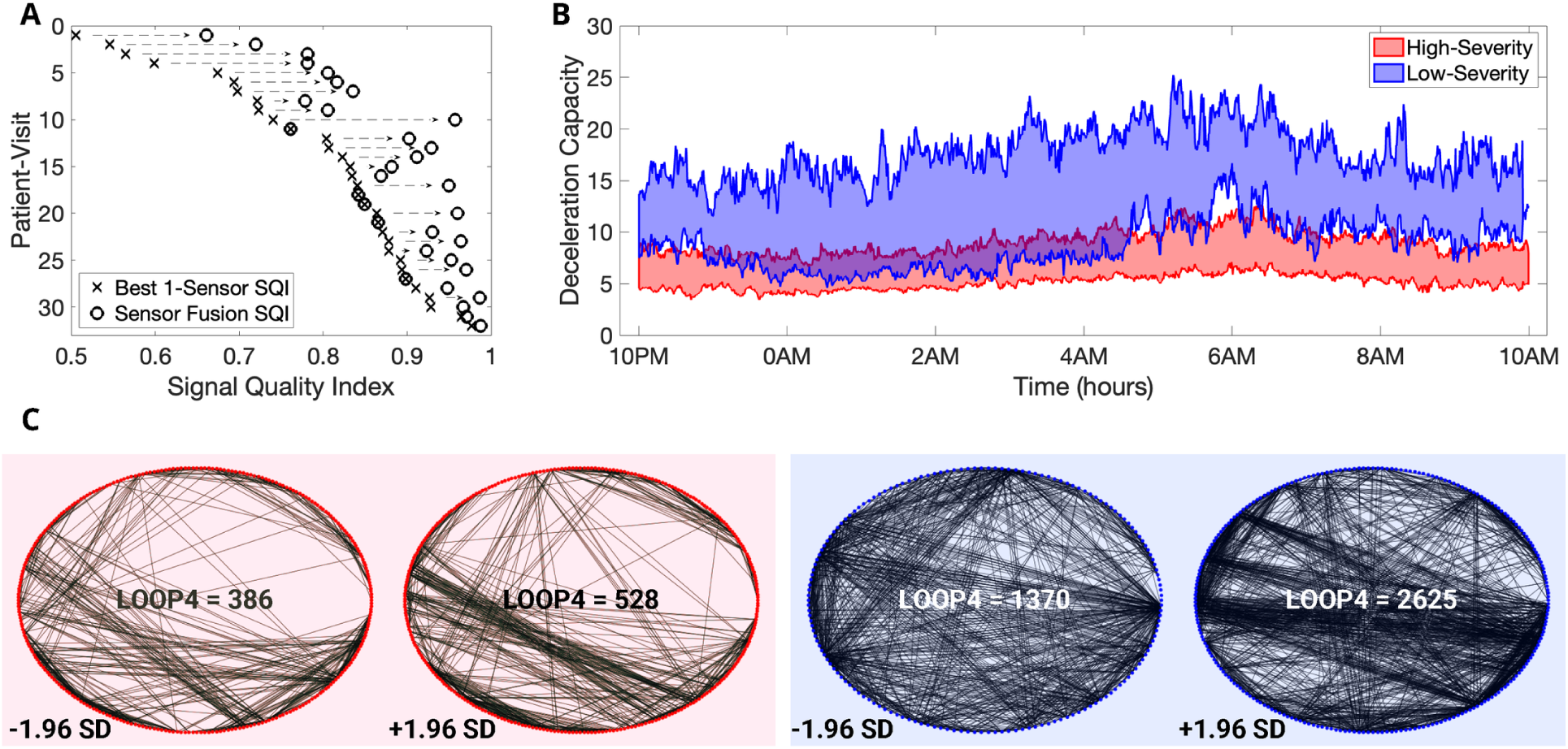
Data imputation techniques combined with first and second-order statistics improve classification performance. (A) An illustration of the effects of novel signal quality index-based ECG data imputation. We show the improvements in the average signal quality index of the electrocardiogram records for all 32 patient-visits. (B) The deceleration capacity (DC) values between 10 PM and 10 AM for the low-severity and high-severity Rett syndrome groups. We show the DC values between the 75^*th*^ and 25^*th*^ percentile for low-severity and high-severity Rett patient-visits. (C) We depict all edges corresponding to 4-loops in the networks constructed from trivariate time series (HR, DC, and Activity count signal) for a high-severity (left-red-panel) and a low-severity (right-blue-panel) patient at the 3^*rd*^ coarse-graining time scale. For each patient, we show two surrogate LOOP4 networks at the following values for LOOP4: (1) 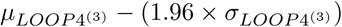, (2) 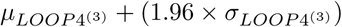. From the image, it is clear that both the number of 4-loops and the variance of the number of 4-loops are smaller for the low-severity patient compared to the high-severity patient.

## Discussion

As of 2022, no clinically meaningful disease-modifying treatments exist for patients with Rett syndrome. We instead rely on multiple therapeutics and symptomatic treatment strategies geared towards managing respiratory ailments, treating seizures, improving gastrointestinal function, and improving motor skills [38]. As new drugs and therapeutics are discovered, the need for objective measures that can be used in clinical trials increases. Neurological disorders, including Rett, suffer from difficult-to-measure symptoms. Most efficacy assessments are based on parent, clinician (and in some cases, patient) interpretation of symptom severity, which by nature introduces bias and often, exemplified by high placebo rates, results in a high noise to signal ratio. Hence, the need to establish objective measurements in patients directly, especially for symptoms that are difficult or impossible to observe, would open the door to evaluate therapeutics in novel ways and has the potential to expedite therapeutic development in multiple ways. Namely, (1) It would reduce bias; (2) It would help reduce clinical trial sample size by reducing the noise to signal ratio; and (3) It would facilitate shorter trial duration by capturing continuous data at home. The measurement of autonomic function could be an early biomarker of therapeutic efficacy and may be particularly relevant for curative therapeutics such as gene therapies that theoretically should improve or restore baseline function. If trials focus on efficacy measures that require learning and implementation (like mobility, communication, and hand use), this may take significantly longer to detect than an autonomic function correction that should not require learning. Thus, in the current study, we attempted to address these unmet needs by capturing physiological (ECG) and body activity (three-axis accelerometer data) from a 20-patient cohort. We chose to regress features extracted from the ECG signal and body activity measurements against the binarized CGI-S to correlate objective measures attained from a wearable biosensor to an overall symptom severity scale. We have shown that the inclusion of multiscale features (MSTE and MSNR) along with HRV-metrics provided a performance improvement of 21% in terms of the AUC (AUC = 0.92) when compared to using HRV-metrics alone (AUC = 0.76). This improvement is attributable to the information embedded in the higher-order interactions between the HR, DC, and activity count signals. While the transfer entropy-based features enabled us to study the 2-dimensional variations between all combinations of HR, DC, and activity count signals in both directions, the network representations-based features enabled us to study the 3-dimensional interactions between these signals. Further, the computations of these features at multiple coarse-graining scales provided a means to analyze the signal modulations occurring due to different physiological phenomena (at different timescales). The coarse-graining scales we used in this study ranged from 1 to 10, which corresponded to a variation of sampling rate from 1*/*30 Hz to 1*/*300 Hz (i.e., from a sample every 30 seconds to a sample every 5 minutes). It allowed us to study the effect of different physiological processes, including respiration, vagal activity, and circadian rhythm, on the interactions between HR, DC, and activity count signals.

Our analysis suggested that the mean value of the DC of the HR signal captured on the BioStamp^®^ nPoint between 10 PM and 10 AM was the most popular feature to classify low-severity Rett syndrome patients from high-severity Rett syndrome patients. It was the most popular feature in all the eight classifier models in which it was used, with a consistent and highest feature popularity score of 1. The computation of DC involved synchronizing the phases of all periodic components of the signal irrespective of their timescales [39]. Thus, the DC captured the overall deceleration of the sinus rhythm due to physiological processes that occurred at different timescales, including the vagal (parasympathetic) activity, the baroreflex, and the circadian rhythm. In Fig 4B, we depicted the variation of the 5 − minute averages of DC between the times of 10 PM and 10 AM for both low-severity and high-severity Rett syndrome patient visits. Apart from the mean value of the DC, we observed the emergence of the variance of transfer entropy at 8^*th*^ coarse-graining scale from (1) Activity count signal to the HR signal and (2) Activity count signal to the DC signal among the most popular features. Further, the variance of the number of loops of size 4 in the network representations at coarse-graining scales 3 and 5 were among the top− 5 most popular features in the best classifier model (AUC = 0.92). Thus, in Fig 4C, we illustrated an example of the network representations for a low-severe and a high-severe Rett patient at the 3^*rd*^, 5^*th*^, and 8^*th*^ coarse-graining scales. In addition to demonstrating the viability of a wearable biosensor to estimate disease severity based on objective measurements directly in patients, another novelty in our work was handling missing data in signals captured using wearables for patient state analysis. We proposed three different techniques for this purpose. The first method of combining data from multiple channels boosted the amount of available data by 13.4% when compared to using a single channel (medial chest) and improved the average SQI by 10%. In Fig 4A, we illustrated this improvement in SQI for each of the 32 patient-visits due to the usage of the novel SQI based ECG imputation. The second method helped impute the activity count data and reduce missingness. The third and final technique of generating stochastic surrogates enabled us to compute the MSTE and MSNR features. It was specifically developed for this study, without which no multiscale analysis could have been performed. We computed the sample mean and sample variance of all MSTE and MSNR metrics across all surrogate representations and used them as features for our classification models. The sample mean is a measure of the samples’ central tendency, and the sample variance is a measure of the spread of the samples. Since our method used a normal distribution to sample the random variables, and a normal distribution can be completely defined by a mean value and variance, it was rational to use the sample mean and variance to characterize the underlying MSTE and MSNR values.

## Conclusion

We developed a machine learning model that utilized features characterizing HRV, body movement, and the interaction between the two to estimate Rett syndrome severity in a group of 20 female Rett patients. For this, we developed a novel stochastic method for biosignal data imputation. We obtained the highest pooled-AUC equal to 0.92 utilizing the feature combination of HRV-metrics, MSTE-features, and MSNR-features. Further, the proposed approach provided us with physio-motor biomarkers that could be used in clinical trials as objective metrics to quantify the improvement in overall patient state. Specifically, the mean DC of the HR signal captured between 10 PM and 10 AM using the BioStamp^®^ nPoint biosensor was the most popular feature with a feature popularity score equal to 1. In conclusion, our study (1) Implemented a novel data imputation technique for physiological signals, (2) Developed a machine learning model to estimate Rett disease severity, and (3) Developed objective measures that characterize the autonomic function in Rett syndrome.

## Data Availability

Data will be posted on PhysioNet (https://physionet.org/) in due course, after de-identification has been verified and permissions for public posting have been secured. Early access to data can be achieved by contacting the authors. The data contains Patient Health Information and hence cannot be publicly shared before de-identification.

## Supporting information

**S1 Fig. The novel feature popularity scores-based visualizations assist in determining the top-5 features for each of the 15 Rett severity classifiers**. We show the normalized feature values for all 32 patient-visits for top-5 most popular features in each subplot. The data points with the red marker correspond to high-severity Rett patient-visits, and the data points with the blue marker correspond to low-severity Rett patient-visits. Further, we provide the feature popularity (*ρ*) scores for these five features. The individual subplots correspond to different feature combinations of the following feature sets: (1) HRV – Heart Rate Variability; (2) Actigraphy; (3) MSTE – Multiscale Transfer Entropy; (4) MSNR – Multiscale Network Representation. (TIF).

